# The Role of Regional and Practice Trial Sites in Distorted Randomized Cancer Trial Enrollment

**DOI:** 10.1101/2025.02.24.25322776

**Authors:** Sachin J. Shah, Christopher R. Manz, Brendan Balthis, Hari S. Raman, Jason Abaluck, Nancy L Keating, Leila Agha

**Author notes:** Address correspondence to: Sachin J Shah, Department of Medicine, Massachusetts General Hospital, 100 Cambridge St, Suite 1600, Boston, MA 02114.

## Abstract

Representative trials are critical to advancing cancer treatment, yet little is known about how geographic siting contributes to non-representative enrollment. Using patient-level data, we determined how the choice of trial-enrolling regions and practices impacts representativeness. We created a SEER-Medicare cohort of people ≥65 years old with lung, breast, pancreatic, or renal cancer (2014-2019). We identified randomized cancer drug trial participants and determined the prevalence of age ≥75, sex, race, ethnicity, and rural residence in the full cohort, trial-enrolling regions, trial-enrolling practices, and trials. The choice of region and practice contributed to >50% of the under-enrollment of Black, Hispanic, and rural patients. Cancer trials enrolled 45% fewer Black patients than expected with proportional representation. Trial recruitment in regions and practices with proportionately fewer Black patients accounted for 27% and 35% of this disparity, respectively. These findings suggest that diversifying cancer trials requires changing the regions and practices referring and enrolling patients.

## INTRODUCTION

The National Academy of Science, the FDA, and professional societies have renewed calls to improve representation in clinical trials, recognizing limited progress over the past 50 years.^1,2^ Under-enrollment of minoritized patients may contribute to low trust, delayed access to therapies, and limited generalizability.^3^ Efforts to diversify trials have focused on improving recruitment within enrolling sites. For example, studies have examined study staff diversity, making an affirmative ask of people underrepresented in research, and active community engagement.^4,5^ Prior work has identified structural barriers within practices that limit recruitment (e.g., trial availability, exclusion criteria), and further upstream efforts have expanded clinical trial access to community oncology practices.^6–8^ Nevertheless, there are scant systematic data on the role of trial siting in representative enrollment. To address this gap, we used patient-level data to determine how the choice of trial-enrolling regions and practices contributes to non-representative enrollment in randomized cancer trials.

## METHODS

### Design, Setting, and Participants

We conducted a longitudinal cohort study of SEER-Medicare participants aged ≥65 years who were diagnosed with lung, breast, pancreatic, or renal cancer between 2014-19. We categorized participants by “disease state,” defined by primary cancer and treatment category, e.g., second-line treatment of advanced-stage non-small cell lung cancer (**Supplementary Table 1**).^9^ We selected these primary sites because they are among the most common primary sites in older adults, and we deemed them most amenable to identifying trial-eligible disease states based on our prior work.^9^ We defined the index date as the date of diagnosis for localized malignancies eligible for neoadjuvant or adjuvant therapies and treatment-line initiation for advanced cancers. To ascertain trial participation and treatment status, we require continuous enrollment in Medicare Part A/B for 12 months before and 6 months after the index date (or until death); we additionally require Part D coverage for patients with renal cancer. We excluded observations missing race, ethnicity, sex, region, or attributable physician practice (**Supplementary Table 2**).

### Region and Practice attribution

Patients were attributed to their hospital referral region of residence at diagnosis. We attributed patients to a clinical practice by taxpayer identification number (TIN).^10^ We identified all carrier and outpatient evaluation & management (E&M) claims within 90 days of the index date. We assigned each patient to the TIN with a plurality of E&M claims among the following specialties: hematology, hematology/oncology, medical oncology, gynecology/oncology **(Supplementary Methods)**. Because some patients travel for care, we attributed practices regardless of whether they were within the patient’s region. Some regions or practices may facilitate access to cancer trials by referring patients to enrolling sites; we captured this functional access and attributed trial enrollment to the patient’s region and practice even if it was not a trial site.

### Race, ethnicity, and rural residence

We used the race and ethnicity categorizations provided by SEER, collected from medical records and supplemented with the NAACCR Hispanic Identification Algorithm.^11^ We defined rural residence as residing in a nonmetropolitan county using Rural-Urban Continuum Codes.^12^

### Trial participation

Patients were potential trial participants if they had a Medicare claim that reported a National Clinical Trial number corresponding to a randomized cancer drug trial, a previously validated measure.^13^ Individuals were considered trial participants if their disease state matched the RCT’s disease state as described on clinicaltrials.gov (**Supplementary Methods** and **Supplementary Figure 1**).

### Analysis

We compared trial participants to non-participants along five sociodemographic characteristics (age ≥75 years, sex, race, ethnicity, and rural residence) across four levels (full cohort, within trial-enrolling regions, within trial-enrolling practices, and within trials). First, we calculated the prevalence of each characteristic among trial participants and in the full cohort. To calculate the prevalence in the full cohort, we used weights to account for the varying trial participation rates across disease states. The difference between the two prevalence estimates represented the “trial gap”: the under or over-representation of a demographic group among trial participants relative to the full cohort.

Next, we calculated the prevalence of sociodemographic characteristics in trial-enrolling regions, weighting each region by the number of trial participants originating from that region for each disease state (i.e., region by disease-state weight). The amount of the trial gap that could be attributed to enrolling regions was calculated as the difference between the full cohort prevalence and the trial-enrolling regions’ prevalence.

Finally, we calculated the prevalence of sociodemographic characteristics in trial-enrolling clinical practices, weighting each practice by the number of trial participants from that practice, region, and disease-state (i.e., practice by region by disease-state weight). The amount of the trial gap that could be attributed to enrolling practices was calculated as the difference between the full cohort prevalence and the trial-enrolling practices’ prevalence. We provide additional details in the **Supplementary Methods**. We calculated, 2-sided, 95% confidence intervals using 1000 bootstrapped samples with replacement. This study was considered exempted research by the Dana-Farber Cancer Institute’s Office for Human Research Studies (#23-610).

## RESULTS

In the study cohort of 206,600 patients, 1,174 participated in 130 cancer drug trials (**Table**). Trials underrepresented Asian and Pacific Islander patients (2.9% trial participants vs. 3.9% full cohort, relative difference -25%, 95% CI -48% to 0%). Under-recruitment of Asian and Pacific Islander patients receiving care at enrolling practices accounted for 72% of this disparity; Asian and Pacific Islander patients were not significantly underrepresented in trial-recruiting regions or practices.

**Table.**
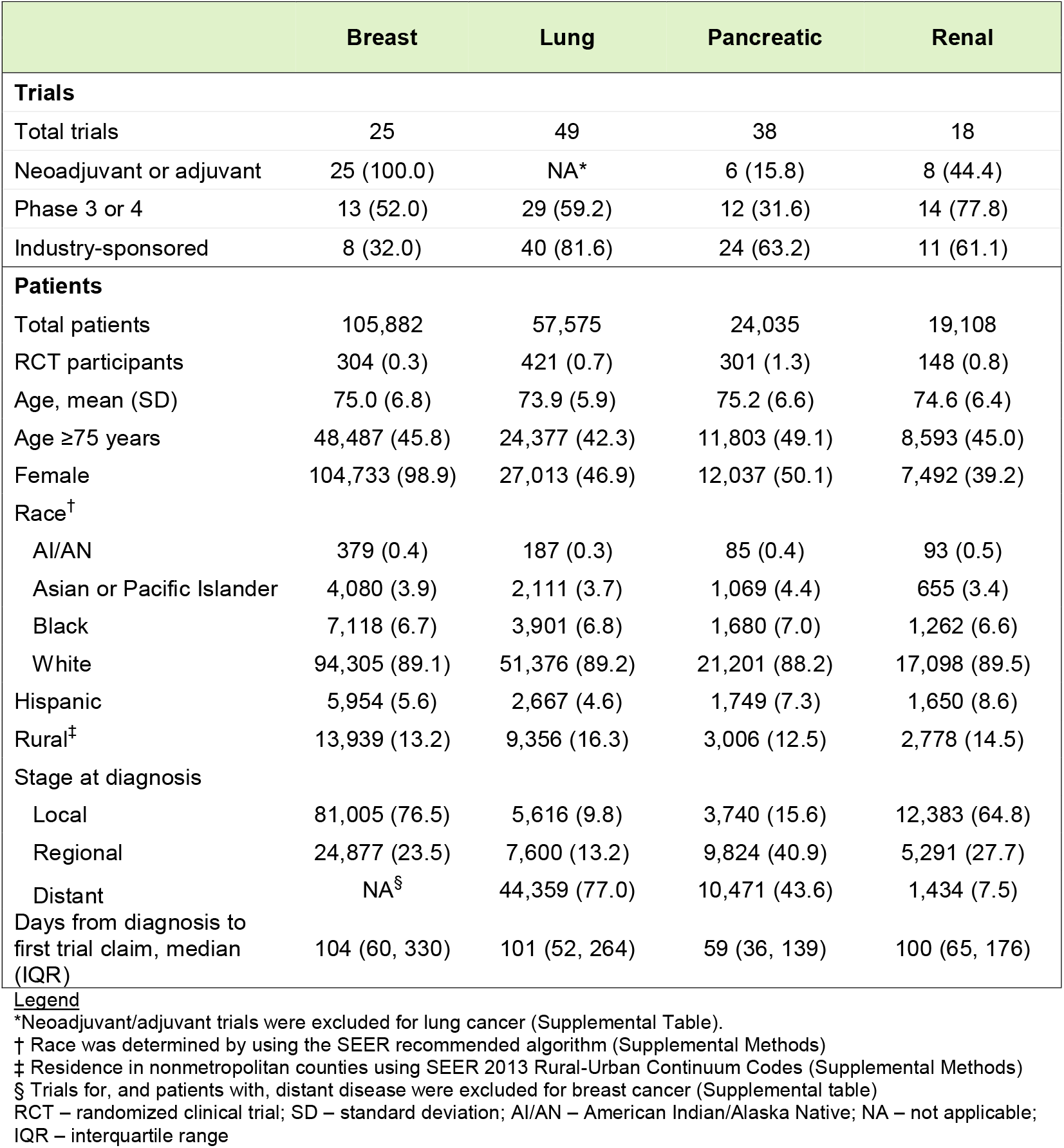
Trial and patient characteristics for SEER-Medicare participants with Breast, Lung, Pancreatic, or Renal cancer from 2014-2019.

|  | Breast | Lung | Pancreatic | Renal |
| --- | --- | --- | --- | --- |
| **Trials** |  |  |  |  |
| Total trials | 25 | 49 | 38 | 18 |
| Neoadjuvant or adjuvant | 25 (100.0) | NA* | 6 (15.8) | 8 (44.4) |
| Phase 3 or 4 | 13 (52.0) | 29 (59.2) | 12 (31.6) | 14 (77.8) |
| Industry-sponsored | 8 (32.0) | 40 (81.6) | 24 (63.2) | 11 (61.1) |
| **Patients** |  |  |  |  |
| Total patients | 105,882 | 57,575 | 24,035 | 19,108 |
| RCT participants | 304 (0.3) | 421 (0.7) | 301 (1.3) | 148 (0.8) |
| Age, mean (SD) | 75.0 (6.8) | 73.9 (5.9) | 75.2 (6.6) | 74.6 (6.4) |
| Age ≥75 years | 48,487 (45.8) | 24,377 (42.3) | 11,803 (49.1) | 8,593 (45.0) |
| Female | 104,733 (98.9) | 27,013 (46.9) | 12,037 (50.1) | 7,492 (39.2) |
| Race<sup>†</sup> |  |  |  |  |
| AI/AN | 379 (0.4) | 187 (0.3) | 85 (0.4) | 93 (0.5) |
| Asian or Pacific Islander | 4,080 (3.9) | 2,111 (3.7) | 1,069 (4.4) | 655 (3.4) |
| Black | 7,118 (6.7) | 3,901 (6.8) | 1,680 (7.0) | 1,262 (6.6) |
| White | 94,305 (89.1) | 51,376 (89.2) | 21,201 (88.2) | 17,098 (89.5) |
| Hispanic | 5,954 (5.6) | 2,667 (4.6) | 1,749 (7.3) | 1,650 (8.6) |
| Rural<sup>‡</sup> | 13,939 (13.2) | 9,356 (16.3) | 3,006 (12.5) | 2,778 (14.5) |
| Stage at diagnosis |  |  |  |  |
| Local | 81,005 (76.5) | 5,616 (9.8) | 3,740 (15.6) | 12,383 (64.8) |
| Regional | 24,877 (23.5) | 7,600 (13.2) | 9,824 (40.9) | 5,291 (27.7) |
| Distant | NA<sup>§</sup> | 44,359 (77.0) | 10,471 (43.6) | 1,434 (7.5) |
| Days from diagnosis to first trial claim, median (IQR) | 104 (60, 330) | 101 (52, 264) | 59 (36, 139) | 100 (65, 176) |

\*Neoadjuvant/adjuvant trials were excluded for lung cancer (Supplemental Table).
† Race was determined by using the SEER recommended algorithm (Supplemental Methods)
‡ Residence in nonmetropolitan counties using SEER 2013 Rural-Urban Continuum Codes (Supplemental Methods)
§ Trials for, and patients with, distant disease were excluded for breast cancer (Supplemental table)
RCT – randomized clinical trial; SD – standard deviation; AI/AN – American Indian/Alaska Native; NA – not applicable;
IQR – interquartile range

Trials enrolled 45% fewer Black patients than would be expected with proportional representation (95% CI -60% to -29%, prevalence 3.7% vs. 6.8%) (**Figure**). Trial recruitment in regions with proportionately fewer Black patients accounted for 27% of this disparity, and the underrepresentation of Black patients at recruiting sites within those regions accounted for a further 34%. Under-recruitment of Black patients within enrolling practices accounted for the remaining 39% of this disparity. White patients were overrepresented in trials.

**Figure:**
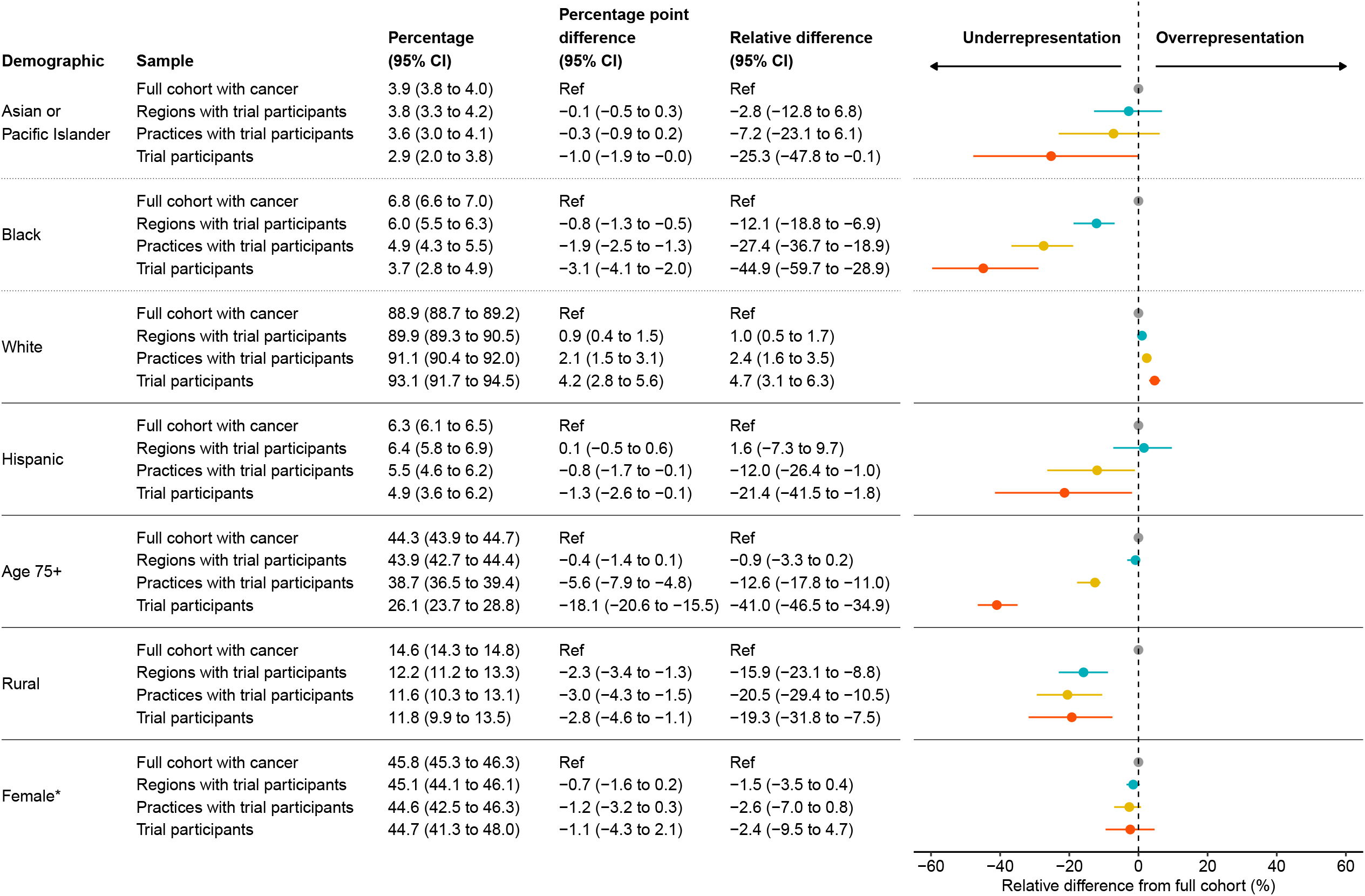
Absolute and relative underrepresentation of patients with cancer at the national, regional, practice, and trial levels weighted by disease state. ^*^ Excludes breast cancer Includes people with breast, lung, pancreatic, or renal cancer in SEER-Medicare diagnosed 2014-2019. The American Indian or Alaska Native patients were not analyzed because fewer than 12 AI/AN patients participated in trials. Disease state was determined by the combination of primary cancer, stage, and treatment line. Absolute values are re-weighted by disease state and level (Supplemental Methods). Confidence intervals were calculated using 1000 bootstrapped samples.

Trials underrepresented Hispanic patients (4.9% vs. 6.3%, relative difference -21%, 95% CI -42% to -2%). Hispanic patients were not significantly underrepresented in trial-recruiting regions. However, trial recruitment in practices with proportionately fewer Hispanic patients accounted for 56% of the disparity. Under-recruitment of Hispanic patients receiving care at enrolling practices accounted for 44% of this disparity.

Patients aged ≥75 years were underrepresented in trials (26.1% vs. 44.3%, relative difference -41%, 95% CI -47% to -35%). Older patients were not significantly underrepresented in trial-recruiting regions. However, trial recruitment in practices with proportionately fewer adults aged ≥75 years accounted for 31% of this disparity. Under-recruitment of adults aged ≥75 years receiving care at enrolling practices accounted for 69% of this disparity.

Trials underrepresented rural residents (11.8% vs. 14.6%, relative difference -19%, 95% CI -32% to -8%). Trial recruitment in regions with proportionately fewer rural patients accounted for 82% of this disparity. Finally, there was no significant underrepresentation of female patients in trials.

## DISCUSSION

This study demonstrated that regions and practices recruiting patients into cancer trials are a structural contributor to the underrepresentation of Black, Hispanic, and rural patients and adults aged ≥ 75 years in cancer trials. More than 50% of the under-representation of Black, Hispanic, and rural patients can be attributed to where trials enroll as opposed to factors within trial enrollment sites (e.g., exclusion criteria, patient preference, bias). These findings imply that expanding trial enrollment sites to regions and clinical practices where Black, Hispanic, and rural patients receive care is essential to closing the enrollment gap. At the same time, substantive literature also highlights that building trust, diverse study staff, and an authentic, longitudinal relationship with communities are essential to diversifying trial enrollment.^1,14^ Thus, expanded trial recruitment sites are likely necessary, but not sufficient to close the trial enrollment gap.

Health policy leaders have called to expand trial access to community practices and advocated for programs like the National Cancer Institute’s Community Oncology Research Program.^15,16^ The study results indicate such efforts have a potentially large impact on improving equitable enrollment through optimal clinical trial siting. Also, in June 2024, the FDA released draft guidance that requires trial sponsors to submit goal patient enrollment disaggregated by age group, sex, and race and ethnicity.^17^ Stated goals must be justified, for example, based on the disease distribution by demographic characteristics. As trial sponsors work to meet these goals, carefully considering the geography of trial sites may be vital.

This study has several important limitations. First, this study was limited because SEER-Medicare does not cover all U.S. patients with cancer. As a result, the study findings may not generalize to younger adults with cancer or states that do not participate in SEER. Second, we used a specific definition of trial participation to ensure the trials were relevant to the matched cohorts with breast, lung, pancreatic, or renal cancer and only included trials with ≥5 SEER-Medicare participants to make the study tractable (**Supplementary Methods 1**). Thus, the trial participation rate in this study does not reflect the overall clinical research participation rate, and the study findings may not generalize to nonrandomized studies (e.g., phase 1 studies, observational studies), to non-drug trials (e.g., surgical trials), nor to other cancer types (e.g., prostate cancer).

This first patient-level analysis improves upon ecological analyses that used area-level racial composition near trial sites.^18–20^ Substantial racial segregation in the health care system cannot be explained by where patients live, so our analysis directly accounts for minoritized patients’ provider choices.^21,22^ This patient-level analysis demonstrated that changing the regions and practices recruiting patients into cancer trials is critical to addressing underrepresentation.

## Data availability

Researchers can apply to SEER to obtain Medicare-linked data.

## Funding

This study was supported by the NIA (K76AG074919). The funders had no role in the design and conduct of the study; collection, management, analysis, and interpretation of the data; preparation, review, or approval of the manuscript; and decision to submit the manuscript for publication.

## Conflict of Interest Disclosure

Dr. Shah reported funding from the National Institute on Aging/National Institutes of Health related to the conduct of this study (noted above). The remaining authors have nothing to disclose.

## Acknowledgments

None

## Supplementary Methods

### 1. Primary outcome: trial participation

Our primary outcome was participation in a randomized drug trial. We started with all NCT numbers found in the Medicare claims of SEER-Medicare participants during the study period (2014-2019) (**Supplementary Figure 1**). Of the NCTs identified, 85% matched a valid clinicaltrials.gov entry and 95% of participants had a valid NCT number. NCTs that could not be matched to clinicaltrials.gov and those that did not correspond to a randomized drug trial were not considered trial participants. We used structured fields from clinicaltrials.gov to identify randomized trials (DesignAllocation) and trials that tested a drug or biological agent (InterventionType). Then, we manually reviewed the set of randomized drug trials with ≥5 SEER-Medicare participants. A physician (HSR) reviewed each clinicaltrials.gov entry to classify the trial’s enrollment criteria vis-à-vis cancer subtype, stage, and treatment line (i.e., disease state). This review excluded trials that were not intended to treat the disease state (e.g., an RCT to prevent anemia) or were not specific to the disease state of interest (e.g., an RCT testing a drug that treats multiple types of solid tumors at once). Thus, individuals were considered trial participants only if the NCT reported in their claims corresponded to a randomized drug trial that matched the patient’s primary cancer subtype, stage, and treatment line. Patients were not considered participants if their NCT-associated Medicare claim preceded their SEER-reported diagnosis date by >90 days.

### 2. Attributing patients to oncology practices

We attributed patients to their plurality oncology practice by taxpayer identification number (TIN) using a modified version of the Keating et al. algorithm.^10^ We identified all Evaluation & Management (E&M) claims in the Carrier and Outpatient files over a 180-day window around each patient’s index date. Since the Outpatient claims do not report provider specialty or TIN numbers, we used each physician’s NPI and merged the physician’s plurality specialty and TIN in the Carrier file to fill in the missing values. We then assigned each patient to the TIN with a plurality of E&M claims among the following specialties: hematology, hematology/oncology, medical oncology, and gynecology/oncology. If there was a tie, we assigned the patient to the TIN with the highest payment value. If a patient had no E&M claims for the above specialties, we broadened the clinical practice set to include family practice, internal medicine, and urology (for patients with renal cancer) claims where the primary diagnosis code was for the cancer of interest.

### 3. Statistical methods

Let *y*_*icrp*_ be an indicator variable that equals 1 if patient *i* belongs to a specific demographic group (e.g., whether the patient is Black). The indices denote patient *i* in disease-state cohort *c*, treated in region *r* and practice *p*.

Among clinical trial participants, we first calculated the unweighted proportion of patients that fell into each sociodemographic group of interest:

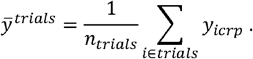

Here, *n*_*trials*_ denotes the number of trial participants, and *i ∈ trials* denotes the subset of patients enrolled in a randomized drug trial.

To compare all patients in our SEER-Medicare cohort to trial participants, we reweighted the national sample to match the proportion of trial participants in each disease state. Because some diseases have higher trial participation rates over our sample period, this reweighting procedure allowed us to compare the demographic composition of trial participants and non-participants, holding the disease state constant. Specifically, we calculated:

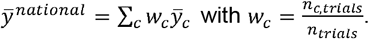

Here, 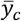denotes the average value of the outcome *y* among all patients (regardless of trial participation status) in disease-state cohort *c*.The weighting variable *w*_*c*_ is defined as the share of all trial participants in disease-state cohort *c*.

Next, to study the composition of patients residing in trial-enrolling regions, we calculated:

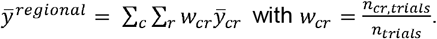

Here, 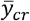 denotes the average value of the outcome *y* among all patients (regardless of trial participation status) in disease-state cohort *c* and region *r*. The weighting variable *w*_*cr*_ is defined as the share of all trial participants in the disease-state cohort *c* and residing in region *r*. Note that *w*_*cr*_ = 0 if there were no trial participants with the patient’s disease-cohort from the patient’s region.

Finally, to study the composition of patients receiving care in trial-enrolling practices, we calculated:

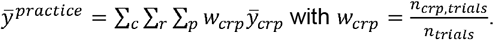

Here, 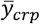 denotes the average value of the outcome *y* among all patients (regardless of trial participation status) in disease-state cohort *c*, region *r*, treated in oncology practice *p*.The weighting variable *w*_*crp*_ is defined as the share of all trial participants that were in disease-state cohort *c*, residing in region *r*, and treated in oncology practice *p*. Note that *w*_*crp*_=0 if there were no trial participants treated in the patient’s oncology practice residing in the same region and with the patient’s disease-state.

In addition to reporting the means defined above, we calculated the difference between the national mean and each of the three other means (region, practice, and trial). 95% confidence intervals were calculated by a bootstrap resampling procedure with 1000 iterations.

**Supplementary Table 1:**
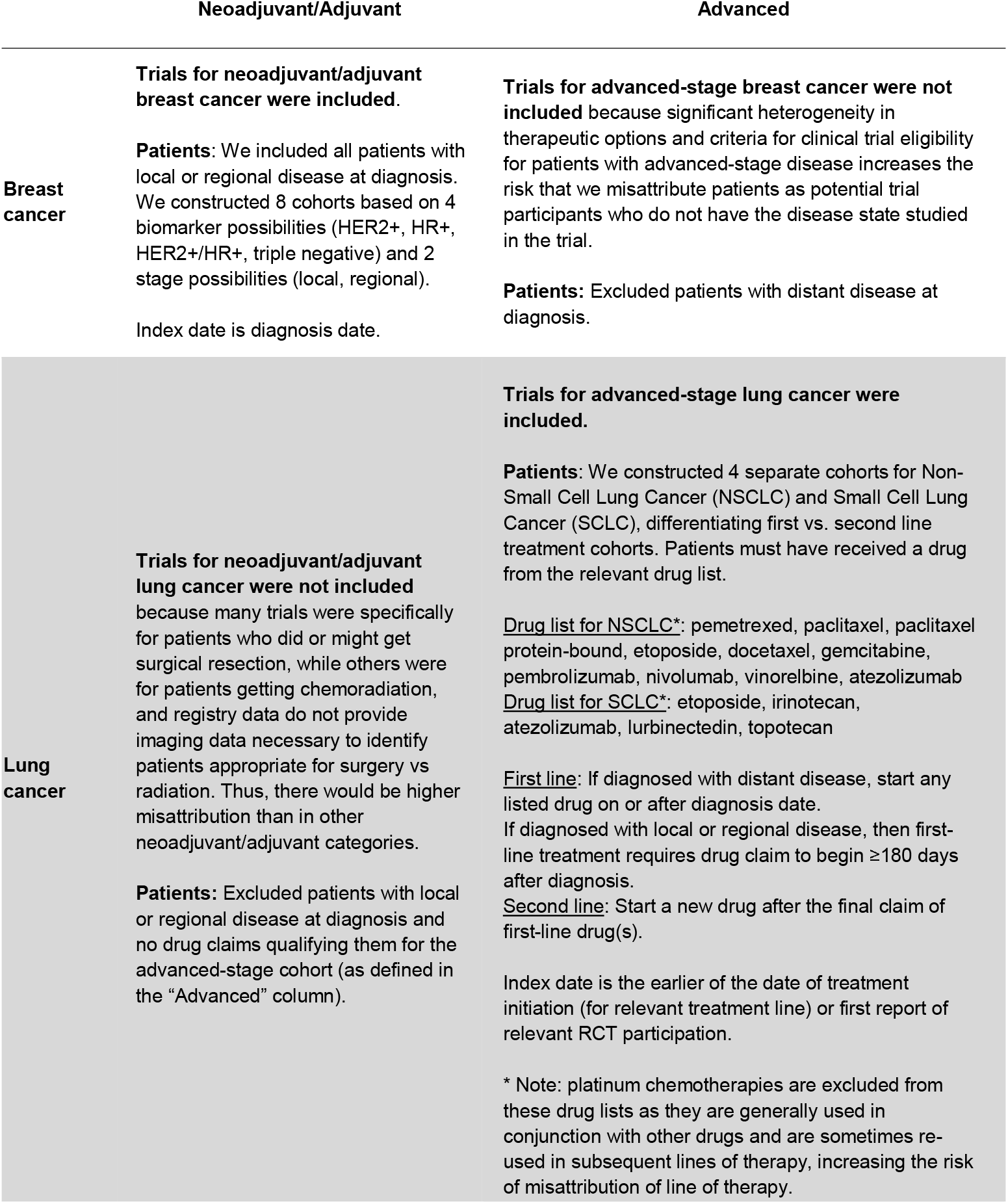
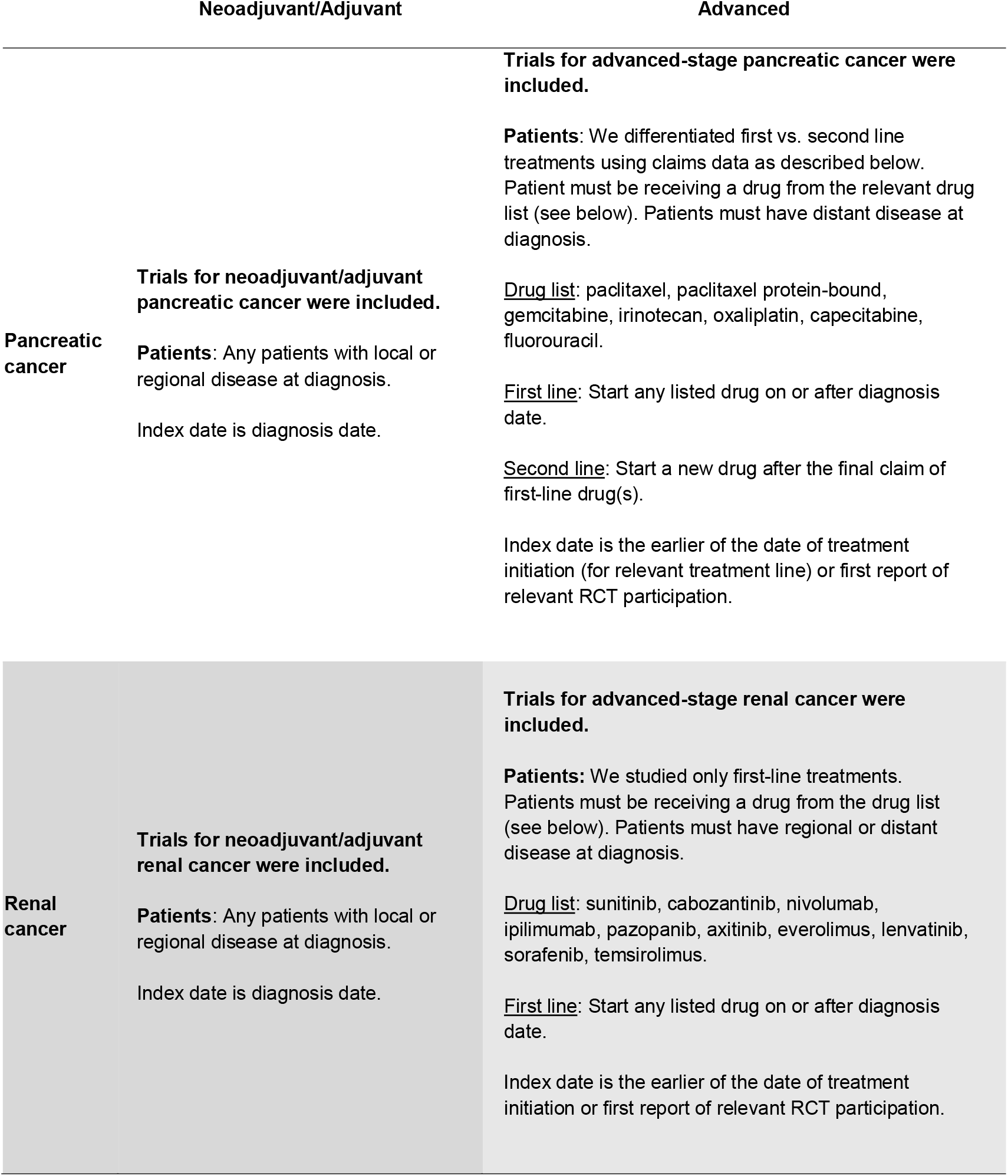
Inclusion and identification of people with trial-eligible cancer drug treatment categories.

**Supplementary Table 2:**
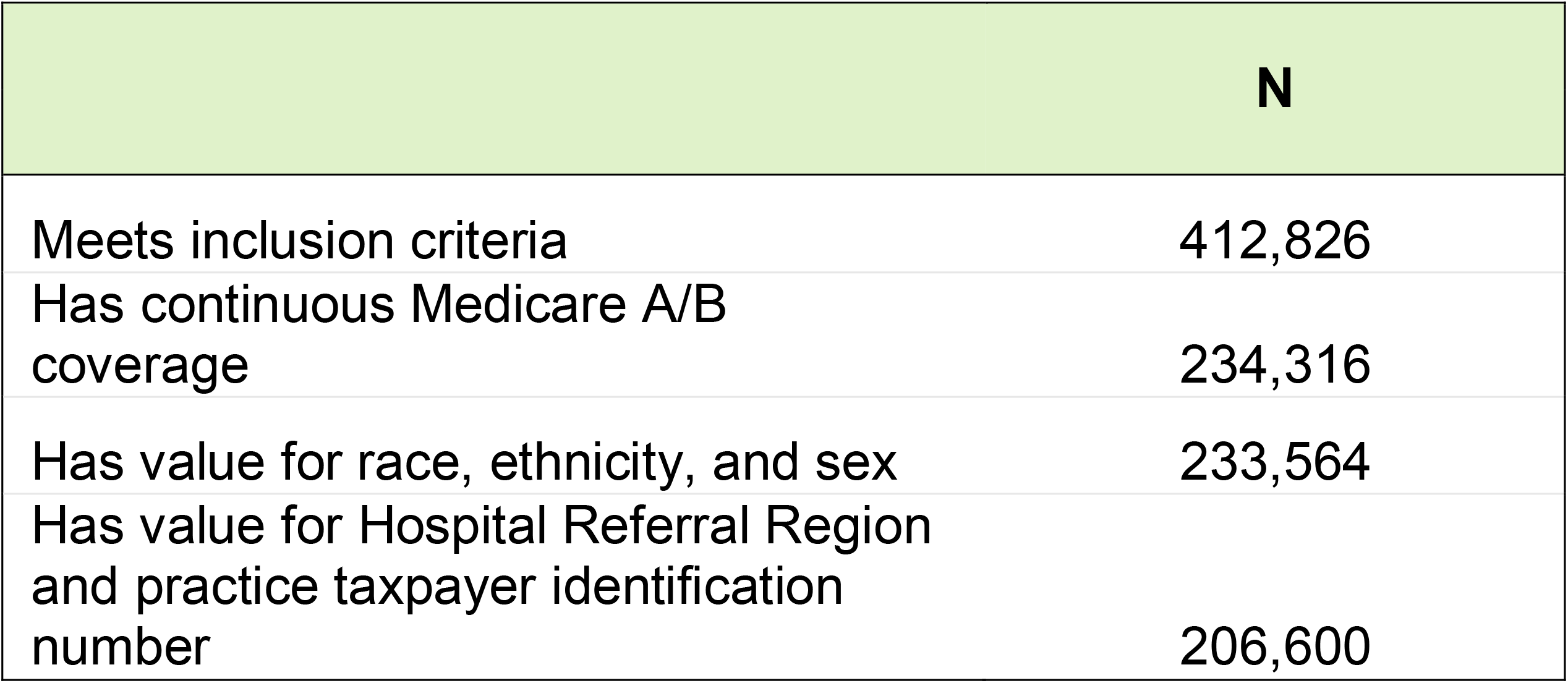
Cohort construction table.

**Supplementary Figure 1:**
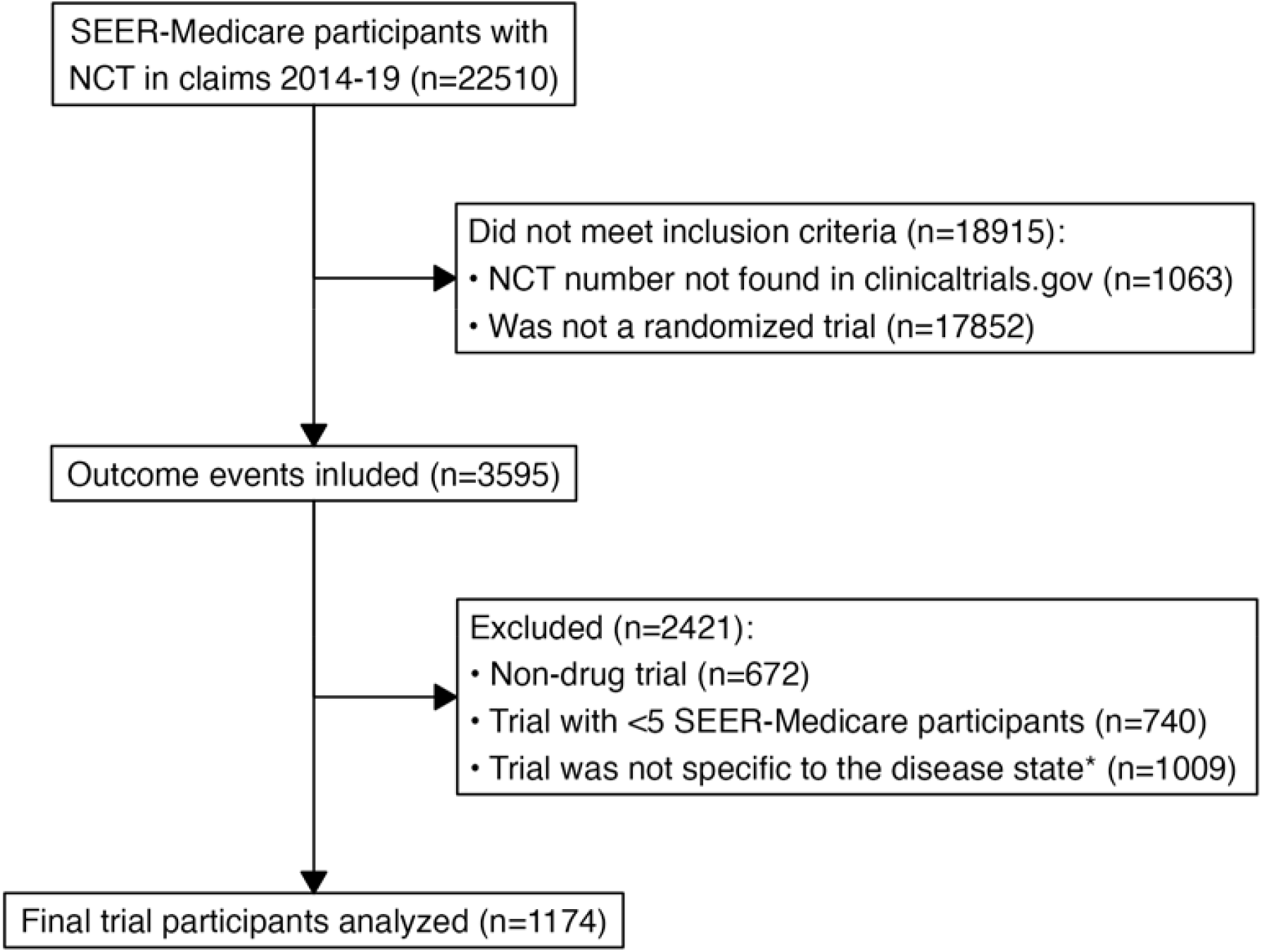
Outcome identification. <u>Legend</u> * We manually reviewed all RCTs to ensure that the cancer drug trial was specific to the referent group’s disease state, i.e., primary cancer type, stage, and line of treatment. Non-cancer directed treatment (e.g., drug trial to prevent chemotherapy-induced anemia) and treatments not directed at the specific primary cancer (e.g., drug treatments directed at multiple solid tumors) were excluded. We made study design choices to increase the specificity of the study population and to make the study tractable. If we were to remove these criteria, the trial participation rate would be 1.74%, approximating the prior literature (1.74% = 3595 / 206,600).^23–26^

## Notes

### Author Declarations

Office for Human Research Studies of the Dana-Farber Cancer Institute's waived ethical approval for this work (protocol number 23-610).

