## supplemental for "The Role of Regional and Practice Trial Sites in Distorted Randomized Cancer Trial Enrollment"

Among clinical trial participants, we first calculated the unweighted proportion of patients that fell into each sociodemographic group of interest:

$$\bar{y}^{trials}=\frac{1}{n_{trials}}\sum_{i\in trials} y_{icrp}.$$

Here, $n_{trials}$ denotes the number of trial participants, and $i\in trials$ denotes the subset of patients enrolled in a randomized drug trial.

To compare all patients in our SEER-Medicare cohort to trial participants, we reweighted the national sample to match the proportion of trial participants in each disease state. Because some diseases have higher trial participation rates over our sample period, this reweighting procedure allowed us to compare the demographic composition of trial participants and non-participants, holding the disease state constant. Specifically, we calculated:

$\bar{y}^{national}=\sum_{c} w_{c}\bar{y}_{c}$with $w_{c}=\frac{n_{c,trials}}{n_{trials}}$.

Here, $\bar{y}_{c}$denotes the average value of the outcome $y$among all patients (regardless of trial participation status) in disease-state cohort $c.$The weighting variable $w_{c}$ is defined as the share of all trial participants in disease-state cohort $c.$

Next, to study the composition of patients residing in trial-enrolling regions, we calculated:

$\bar{y}^{regional}= \sum_{c} \sum_{r} w_{cr}\bar{y}_{cr}$with $w_{cr}=\frac{n_{cr,trials}}{n_{trials}}$.

Here, $\bar{y}_{cr}$ denotes the average value of the outcome $y$among all patients (regardless of trial participation status) in disease-state cohort $c$ and region $r$. The weighting variable $w_{cr}$ is defined as the share of all trial participants in the disease-state cohort $c$ and residing in region $r.$ Note that $w_{cr}=0$ if there were no trial participants with the patient’s disease-cohort from the patient’s region.

Finally, to study the composition of patients receiving care in trial-enrolling practices, we calculated:

$\bar{y}^{practice}=\sum_{c} \sum_{r} \sum_{p} w_{crp}\bar{y}_{crp}$with $w_{crp}=\frac{n_{crp,trials}}{n_{trials}}$.

Here, $\bar{y}_{crp}$denotes the average value of the outcome $y$among all patients (regardless of trial participation status) in disease-state cohort $c$, region $r$, treated in oncology practice $p.$ The weighting variable $w_{crp}$ is defined as the share of all trial participants that were in disease-state cohort $c$, residing in region $r,$ and treated in oncology practice $p.$ Note that $w_{crp}=0$ if there were no trial participants treated in the patient’s oncology practice residing in the same region and with the patient’s disease-state.

In addition to reporting the means defined above, we calculated the difference between the national mean and each of the three other means (region, practice, and trial). 95% confidence intervals were calculated by a bootstrap resampling procedure with 1000 iterations.

### **Supplementary Table 1: Inclusion and identification of people with trial-eligible cancer drug treatment categories**

|  | **Neoadjuvant/Adjuvant** | **Advanced** |
| --- | --- | --- |
| **Breast cancer** | **Trials for neoadjuvant/adjuvant breast cancer were included**.  **Patients**: We included all patients with local or regional disease at diagnosis. We constructed 8 cohorts based on 4 biomarker possibilities (HER2+, HR+, HER2+/HR+, triple negative) and 2 stage possibilities (local, regional).  Index date is diagnosis date. | **Trials for advanced-stage breast cancer were not included** because significant heterogeneity in therapeutic options and criteria for clinical trial eligibility for patients with advanced-stage disease increases the risk that we misattribute patients as potential trial participants who do not have the disease state studied in the trial.  **Patients:** Excluded patients with distant disease at diagnosis. |
| **Lung cancer** | **Trials for neoadjuvant/adjuvant lung cancer were not included** because many trials were specifically for patients who did or might get surgical resection, while others were for patients getting chemoradiation, and registry data do not provide imaging data necessary to identify patients appropriate for surgery vs radiation. Thus, there would be higher misattribution than in other neoadjuvant/adjuvant categories.  **Patients:** Excluded patients with local or regional disease at diagnosis and no drug claims qualifying them for the advanced-stage cohort (as defined in the “Advanced” column). | **Trials for advanced-stage lung cancer were included.**  **Patients**: We constructed 4 separate cohorts for Non-Small Cell Lung Cancer (NSCLC) and Small Cell Lung Cancer (SCLC), differentiating first vs. second line treatment cohorts. Patients must have received a drug from the relevant drug list.  Drug list for NSCLC*: pemetrexed, paclitaxel, paclitaxel protein-bound, etoposide, docetaxel, gemcitabine, pembrolizumab, nivolumab, vinorelbine, atezolizumab Drug list for SCLC*: etoposide, irinotecan, atezolizumab, lurbinectedin, topotecan  First line: If diagnosed with distant disease, start any listed drug on or after diagnosis date. If diagnosed with local or regional disease, then first-line treatment requires drug claim to begin ≥180 days after diagnosis.  Second line: Start a new drug after the final claim of first-line drug(s).  Index date is the earlier of the date of treatment initiation (for relevant treatment line) or first report of relevant RCT participation.  * Note: platinum chemotherapies are excluded from these drug lists as they are generally used in conjunction with other drugs and are sometimes re-used in subsequent lines of therapy, increasing the risk of misattribution of line of therapy. |
| **Pancreatic cancer** | **Trials for neoadjuvant/adjuvant pancreatic cancer were included.**  **Patients**: Any patients with local or regional disease at diagnosis.  Index date is diagnosis date. | **Trials for advanced-stage pancreatic cancer were included.**  **Patients**: We differentiated first vs. second line treatments using claims data as described below. Patient must be receiving a drug from the relevant drug list (see below). Patients must have distant disease at diagnosis.  Drug list: paclitaxel, paclitaxel protein-bound, gemcitabine, irinotecan, oxaliplatin, capecitabine, fluorouracil.  First line: Start any listed drug on or after diagnosis date.  Second line: Start a new drug after the final claim of first-line drug(s).  Index date is the earlier of the date of treatment initiation (for relevant treatment line) or first report of relevant RCT participation. |
| **Renal cancer** | **Trials for neoadjuvant/adjuvant renal cancer were included.**  **Patients**: Any patients with local or regional disease at diagnosis.  Index date is diagnosis date. | **Trials for advanced-stage renal cancer were included.**  **Patients:** We studied only first-line treatments. Patients must be receiving a drug from the drug list (see below). Patients must have regional or distant disease at diagnosis.  Drug list: sunitinib, cabozantinib, nivolumab, ipilimumab, pazopanib, axitinib, everolimus, lenvatinib, sorafenib, temsirolimus.  First line: Start any listed drug on or after diagnosis date.  Index date is the earlier of the date of treatment initiation or first report of relevant RCT participation. |

### **Supplementary Table 2: Cohort construction table**

|  | **N** |
| --- | --- |
| Meets inclusion criteria | 412,826 |
| Has continuous Medicare A/B coverage | 234,316 |
| Has value for race, ethnicity, and sex | 233,564 |
| Has value for Hospital Referral Region and practice taxpayer identification number | 206,600 |

### **Supplementary Figure 1: Outcome Identification**

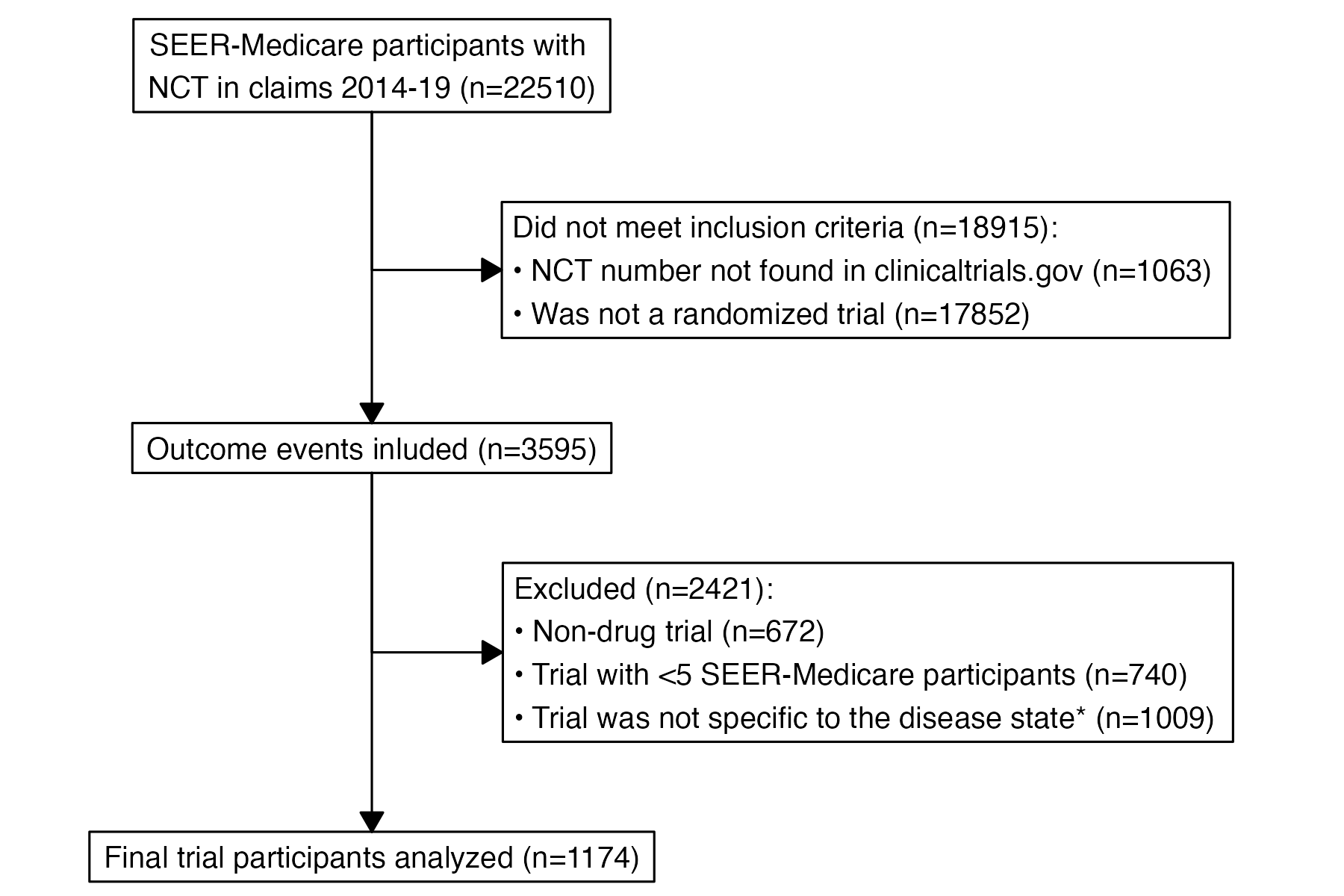

Legend

* We manually reviewed all RCTs to ensure that the cancer drug trial was specific to the referent group’s disease state, i.e., primary cancer type, stage, and line of treatment. Non-cancer directed treatment (e.g., drug trial to prevent chemotherapy-induced anemia) and treatments not directed at the specific primary cancer (e.g., drug treatments directed at multiple solid tumors) were excluded. We made study design choices to increase the specificity of the study population and to make the study tractable. If we were to remove these criteria, the trial participation rate would be 1.74%, approximating the prior literature (1.74% = 3595 / 206,600).^23–26^
